# Associations of perceived discrimination with health outcomes and health disparities in the *All of Us* cohort

**DOI:** 10.1101/2024.10.11.24315343

**Authors:** Vincent Lam, Sonali Gupta, I. King Jordan, Leonardo Mariño-Ramírez

**Author notes:** Correspondence: Leonardo Mariño-Ramírez 11545 Rockville Pike Building 11545 Rockville Pike 2WF Room C14 Rockville, MD 20818.

## Abstract

**Objective:** The goal of this study was to investigate the association of perceived discrimination with health outcomes and disparities.

**Materials and Methods:** The study cohort consists of 60,180 participants from the four largest SIRE groups in the *All of Us* Research Program participant body: Asian (1,291), Black (4,726), Hispanic (5,336), and White (48,827). A perceived discrimination index (PDI) was derived from participant responses to the “Social Determinants of Health” survey, and the *All of Us* Researcher Workbench was used to analyze associations and mediation effects of PDI and self-identified race and ethnicity (SIRE) with 1,755 diseases.

**Results:** The Black SIRE group has the greatest median PDI, followed by the Asian, Hispanic, and White groups. The Black SIRE group shows the greatest number of diseases with elevated risk relative to the White reference group, followed by the Hispanic and Asian groups. PDI was found to be positively and significantly associated with 489 out of 1,755 (27.86%) diseases. ‘Mental Disorders’ is the disease category with the greatest proportion of diseases positively and significantly associated with PDI: 59 out of 72 (81.94%) diseases. Mediation analysis showed that PDI mediates 69 out of 351 (19.66%) Black-White disease disparities.

**Discussion:** Perceived discrimination is significantly associated with risk for numerous diseases and mediates Black-White disease disparities in the *All of Us* participant cohort.

**Conclusion:** This work highlights the role of discrimination as an important social determinant of health and provides a means by which it can be quantified and modeled on the *All of Us* platform.

## INTRODUCTION

The expanding focus on the social determinants of health (SDOH) in health research is reflected in the growing literature on SDOH in the United States (US) and beyond [1]. The World Health Organization defines social determinants of health as “the non-medical factors that influence health outcomes”. SDOH can include income, education, healthcare access, housing stability, and social support, among other factors related to the conditions and environments under which individuals live and work [2]. SDOH are estimated to influence up to 60% of health outcomes [3], with adverse SDOH exposures being associated poorer health outcomes. For instance, infant mortality is significantly higher in rural and poorer areas [4]. Furthermore, mortality and prevalence rates for specific diseases such as COVID-19, cancer, diabetes, and cardiovascular disease are greater for those of lower socioeconomic status (SES) and for those residing in deprived neighborhoods [5–8].

Discrimination—the unequal treatment of individuals on the grounds of physical characteristics or membership to particular demographic groups—is a SDOH with negative effects on health outcomes. Such negative effects often manifest as mental health disorders, as individuals who experience discrimination demonstrate greater risk of ailments such as psychosis, suicidal ideation, and depression [9–11]. Though to a generally lesser extent, discrimination is also a predictor for poorer physical health [12]. One potential mechanism through which discrimination may worsen physical health outcomes is increasing the likelihood of engaging in behaviors that negatively impact health, such as substance abuse [13–15]. Discrimination may also impact physical health through more direct routes, such as by increasing red blood cell oxidative stress and affecting cortisol levels in the body [16 17].

Discrimination may contribute to health disparities. The National Institute on Minority Health and Health Disparities (NIMHD) defines health disparities as “health difference[s] that adversely affect disadvantaged populations”. Health disparities are widespread in the United States, with minority racial groups faring worse than the majority White population across several health measures, including life expectancy, disease prevalence, and perceived health [18 19]. Racial discrimination may contribute to these disparities by driving racial differences in SES, a known driver of health disparities [20]. Racial discrimination may also result in minority racial groups receiving poorer quality of health care compared to White patients [21]. Furthermore, as racial minority groups in the United States generally report greater levels of perceived discrimination than their White counterparts, such groups may be more vulnerable to the negative health effects that accompany discrimination [22 23]. As such, discrimination may help explain the disproportionately negative health outcomes experienced by disadvantaged groups. We hypothesized that a connection between discrimination and health disparities may be uncovered using the *All of Us* Research Program’s (abbreviated as *All of Us* hereafter) data on perceived discrimination, demography, and health outcomes.

*All of Us* is a federal initiative launched in 2015 with the goal of advancing precision and health equity through the collection of genetic, health, and demographic data from over one million American volunteers [24]. Among the program’s priorities is the recruitment of individuals from demographic groups that have been historically underrepresented in biomedical research. The resulting diversity of its participant body, coupled with the program’s wealth of data on health status and discrimination, make it a compelling platform from which to study the confluence of discrimination and health disparities.

The data on perceived discrimination present on the *All of Us* platform takes the form of participant responses to a series of survey questions. The first aim of this study was to use these survey responses to derive a quantitative measure of perceived discrimination that can be associated with health outcomes. The second aim of this study was to use this derived metric to assess how perceived discrimination is associated with health outcomes and health disparities across the *All of Us* participant cohort. The metric produced in this study can aid future research on discrimination in the *All of Us* cohort by providing a means by which perceived discrimination can be quantified and modeled. The accompanying analyses elucidate the role of perceived discrimination in driving health disparities and provide a broad picture of how discrimination affects health and disease.

## MATERIALS AND METHODS

### Study cohort

The cohort for this study was assembled and analyzed using participant data obtained from the cloud-based *All of Us* Researcher Workbench. Volunteers can enroll in the program online through JoinAllofUs.org or through a participating healthcare provider. Enrollment is restricted to individuals 18 years of age or older and to those residing in the US or in a US territory. Individuals who are incarcerated or unable to provide consent are not eligible to enroll in the program.

*All of Us* participant data were taken from the Registered Tier Dataset v7 (curated version R2022Q4R9), from which participant demographic, electronic health record (EHR), and survey data were obtained. Enrollment began on May 31^st^, 2017. The cutoff date for v7 of the Registered Tier Dataset is July 1^st^, 2022. Extracted demographic data consisted of participant self-identified race and ethnicity (SIRE), date of birth, and sex at birth. International Classification of Diseases codes (ICD-9-CM and ICD-10-CM) were extracted from participant EHR data and mapped to 1,755 disease phecodes to designate participants as either a case or control for each disease [25].

The study cohort was restricted to participants who had SIRE data, were assigned either male or female at birth, as to facilitate the use of sex as a covariate in regression analyses, who have EHR data available, and who have provided responses to survey questions on perceived discrimination.

### Race and ethnicity

In the *All of Us* survey titled “The Basics”, participants are asked to select one or more of seven racial and ethnic categories that they most closely identify as: (1) American Indian or Alaska Native, (2) Asian, (3) Black, (4) Hispanic or Latino, (5) Middle Eastern or North African, (6) Native Hawaiian or Pacific Islander, and (7) White. Participants are also given the option to respond with “None of these fully describe me” or “Prefer not to answer.” The *All of Us* Researcher Workbench codes these data as self-identified race and ethnicity categories, following the US Office of Management and Budget Standards. SIRE data are currently not available for those who selected the ‘American Indian or Alaska Native’ category. To maximize statistical power, the study cohort was limited to the four largest SIRE groups in the *All of Us* participant body. Asian, Black, and White participants were defined as those who selected the corresponding category as their sole category, and Hispanic participants were defined as individuals who selected ‘Hispanic or Latino’ as either one of or their only category, consistent with OMB standards.

### Quantifying perceived discrimination

A metric for quantifying the discrimination that *All of Us* participants perceive was derived from participant responses to the program’s “Social Determinants of Health” survey. In the section of the survey centered on discrimination, participants are asked to state the frequency with which they experience nine different forms of discrimination, corresponding to nine different questions. The questions read, “In your day-to-day life, how often do any of these happen to you?”: (1) You are treated with less courtesy than other people are, (2) You are treated with less respect than other people are, (3) You receive poorer service than other people at restaurants or stores, (4) People act as if they think you are not smart, (5) People act as if they are afraid of you, (6) People act as if they think you are dishonest, (7) People act as if they’re better than you are, (8) You are called names or insulted, and (9) You are threatened or harassed. Participants may respond with one of six options corresponding to ascending levels of frequency: (1) Never, (2) Less than once a year, (3) A few times a year, (4) A few times a month, (5) At least once a week, and (6) Almost every day. Furthermore, participants are asked to cite an attribution for these experiences: “What do you think is the main reason for these experiences”. This survey section is adapted from the Everyday Discrimination Scale, which was developed by Williams and colleagues to measure perceived discrimination [26 27].

These six possible responses were coded as ordinal values ranging from 0 to 5, with higher values corresponding to a higher frequency of perceived discrimination. A matrix consisting of participant ordinal responses to each of the nine survey questions was converted to a polychoric correlation matrix using the *polychoric* function from version 2.4.1 of the *psych* package in R version 4.3.1 [28]. The *eigen* function was used to perform principal component analysis on the polychoric correlation matrix. A new matrix was generated consisting of eigenvectors (principal components) arranged in decreasing order of their respective eigenvalues. The first principal component was min-max normalized and taken used as metric for each participant’s perceived discrimination, termed the perceived discrimination index (PDI).

### Statistical analysis

All statistical analyses in this study were performed on R version 4.3.1. Logistic regression models were constructed using the *stats* package’s *glm* function. Logistic regression models were used to model disease status (case=1, control=0) as a function of PDI, PDI as a function of SIRE, and disease status as a function of different combinations of SIRE and PDI. When modeling disease status, participant age and sex at birth were used as covariates to mitigate bias produced by the overrepresentation of older and female volunteers in the *All of Us* participant body.

The contributions of perceived discrimination to disease disparities across SIRE groups were evaluated using two sets of logistic regression models. These analyses were performed on diseases for which a significant and positive association between disease status and membership to either the Asian, Black, or Hispanic SIRE groups were observed. In an unadjusted set of models, disease status was modeled as a function of SIRE with age and sex at birth as covariates. In an adjusted set of models, disease status was modeled as a function of SIRE with age, sex at birth, and PDI as covariates. For a given disease for a particular SIRE group, the extent to which perceived discrimination contributes to disparities in disease prevalence between the SIRE group and the White SIRE group was quantified using percent attenuation, PDI_attenuation_. This value represents the percent decrease in the coefficient for the SIRE group in predicting case status following adjustment for PDI:

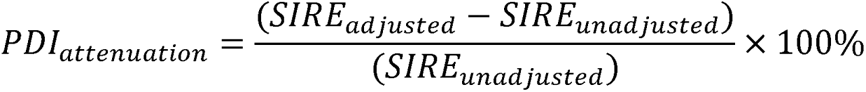

The contribution of perceived discrimination to disparities in cross-SIRE disease burden was also evaluated through mediation analysis. Mediation analysis was performed using version 10.7-1 of the *mma* package. In such an analysis, the total effects of an exposure on a disease outcome is decomposed into the direct effect of the exposure and the indirect effect of the exposure that is mediated by a second variable. In this study, membership to the Black SIRE group was identified as the exposure and PDI as the mediator contributing to disease case status.

## RESULTS

### Study cohort

Data were available for a total of 413,457 participants in the *All of Us* Registered Tier Dataset v7. The study cohort was limited to participants who had both demographic and EHR data available and who provided responses to questions regarding discrimination in the “Social Determinants of Health” survey (Supplementary Figure 1). The study cohort was further limited to participants belonging to the Asian, Black, Hispanic, and White SIRE groups and who were assigned either male or female at birth. These restrictions were employed to attain adequate statistical power and to allow adjustment by sex at birth. The final study cohort featured a total of 60,180 participants, whose mean age is 60.66 years and 65.00% were assigned female at birth (Supplementary Table S1).

### Perceived discrimination index (PDI)

A perceived discrimination index (PDI) for *All of Us* participants was derived from participant responses to nine questions regarding discrimination in the “Social Determinants of Health” survey (Supplementary Table S2). Individual values for PDI were computed for 107,723 participants, of whom 60,180 met the study inclusion criteria. The PDI is a composite metric of discrimination that *All of Us* participants perceive, incorporating frequencies that participants report for each of nine questions describing experiences of discrimination. The questions capture different forms of discrimination that an individual may experience. Reported frequencies were coded as ordinal values, with higher values representing higher frequencies. Polychoric correlation (ρ) tests performed on each pair of questions reveal that reported frequencies for these questions are highly and positively correlated. Correlation scores range from ρ=0.49 for questions 2 and 5 to ρ=0.91 for questions 3 and 4 (Figure 1A). The mean correlation score for all pairwise correlations was ρ=0.63.

**Figure 1.**
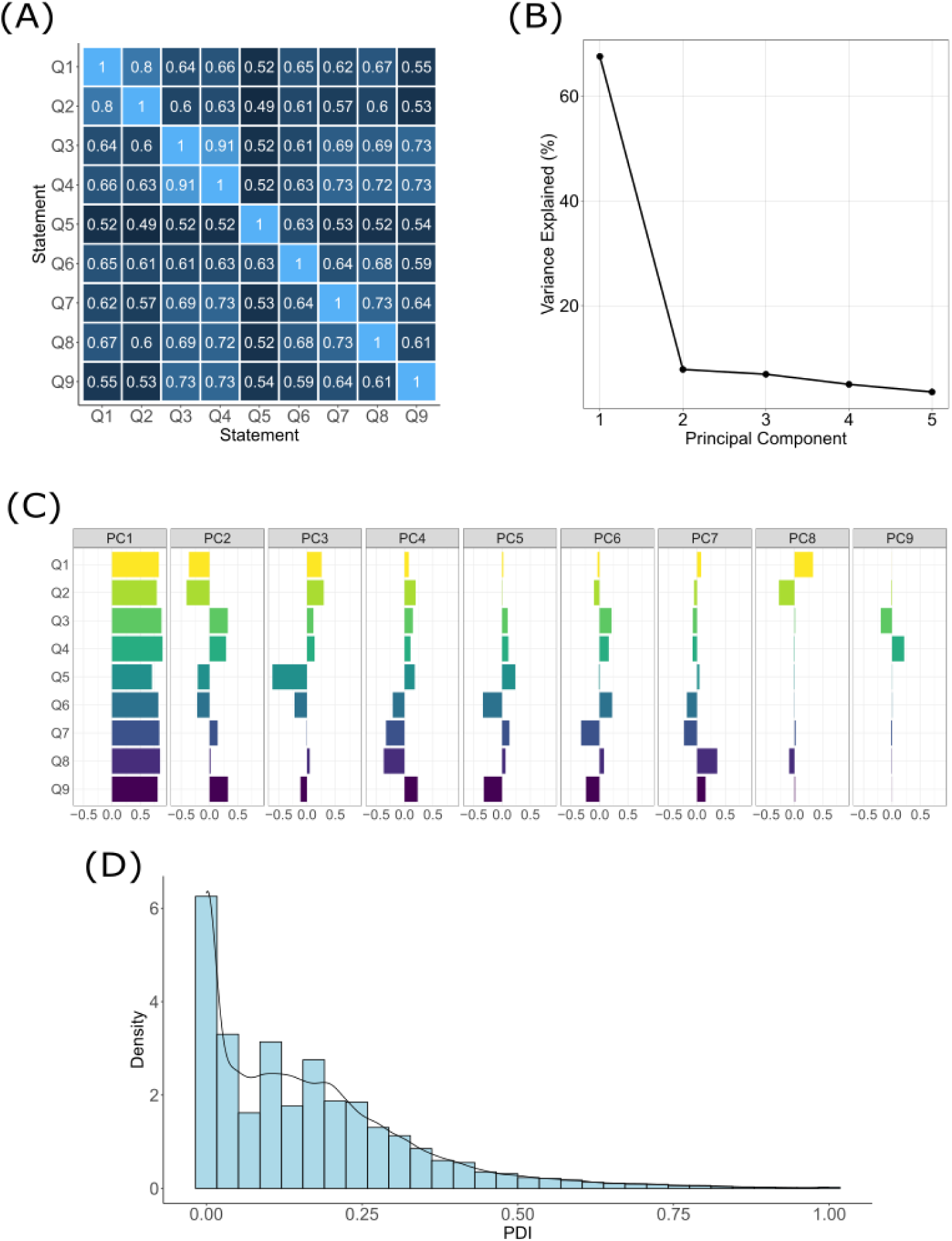
Development of the perceived discrimination index (PDI). (A) Pairwise polychoric correlation scores for ordinally-coded responses to the nine discrimination-related questions from the “Social Determinants of Health” survey. (B) The proportion of variance in participant responses to the nine discrimination-related questions explained (y-axis) by each of the first five principal components produced through principal component analysis of the polychoric correlation matrix (x-axis). (C) Principal component loadings for each of the nine discrimination-related questions (y-axis). (D) Distribution of participant PDI values (x-axis) for the study cohort.

Principal component analysis was applied to the ordinal-coded frequencies to generate single, participant-specific values that capture the variation in reported frequencies across the nine questions. The first principal component (PC1) explains 67.54% of the variance in participants’ reported frequencies, followed by 7.82% for PC2 and 6.90% for PC3 (Figure 1B). The variable loadings for PC1 are all positive, with higher loading values corresponding to greater degrees of perceived discrimination. The greatest loading value observed was 0.89 for question and the lowest loading value observed was 0.70 for question 5 (Figure 1C). The mean loading value across all nine questions was 0.82. As PC1 explained the greatest amount of variance in reported frequencies and demonstrated consistently positive variable loadings, values for PC1 were used to calculate PDI. Min-max normalization was applied to PC1 to produce a continuum of PDI values ranging from 0 to 1, with higher values corresponding to greater degrees of perceived discrimination (Figure 1D).

Differences in the degree of discrimination perceived by individuals belonging to different SIRE groups was assessed through modeling PDI as a function of SIRE, with participant age and sex at birth as covariates. Using White as the reference SIRE group, positive and significant associations were observed between PDI and the Asian (β=1.84e-2, p=1.61e-5) and Black (β=8.74e-2, p=1.91 e−312) SIRE groups (Supplementary Table S3). These associations remain when covariates are removed (Supplementary Table S4). No significant associations were observed between PDI and the Hispanic SIRE group. The patterning in these model coefficient values is reflected in the median PDI values for the four SIRE groups. The Black SIRE group has the highest median PDI of 0.20, followed by the Asian, Hispanic, and White SIRE groups, with scores of 0.18, 0.14, and 0.11, respectively (Figure 2). The reasons that participants gave for experiencing discrimination that were most significantly and positively associated with PDI values are “Some Other Aspect of Your Physical Appearance” (β=7.33e-2, p=5.76e-195), followed by “Your Weight” (β=5.75e-2, p=1.87e-150), and “Your Race” (β=5.08e-2, p=1.49e-128) (Supplementary Table S5).

**Figure 2.**
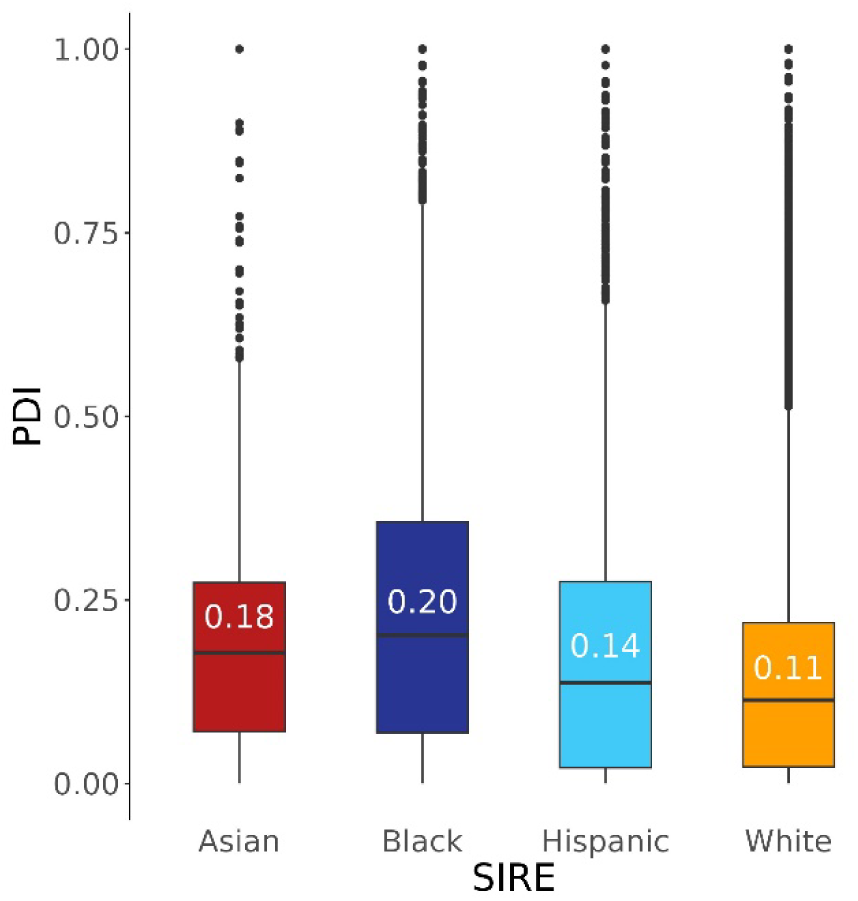
Distribution of perceived discrimination index (PDI) scores for participant self-identified race ethnicity (SIRE) groups. Boxplots show the median, interquartile ranges, and outliers, with median values included.

### Perceived discrimination and health outcomes

To assess the association between perceived discrimination and health outcomes across the *All of Us* participant body, logistic regression models were used to model disease status (case=1, control=0) as a function of PDI for a total of 1,755 diseases (Figure 3A). Participant age and sex at birth were included as covariates in all models, as prevalence for a number of diseases are known to differ across sex and age groups. Of the 1,755 diseases modeled, 489 (27.86%) were positively and significantly (Bonferroni adjusted p<2.85×10^-5^) associated with PDI and 20 (1.14%) were negatively and significantly associated with PDI. ‘Mental Disorders’ is the disease category with the largest percent of diseases significantly and positively associated with PDI, with a total of 59 out of 72 (81.94%) PDI associated diseases, followed by ‘Neurological Disorders’, which has a total of 43 out of 82 associated diseases (52.43%) (Figure 3B). The disease most strongly and positively associated with PDI is posttraumatic stress disorder (β=3.51, p=1.19e-231), followed by major depressive disorder (β=1.94, p=1.41e-195) and mood disorders (β=1.94, p=7.66e-169) (Table 1). Similar results are achieved when perceived discrimination is represented as a categorical variable, such as in previous studies conducted by Forde and colleagues (Supplementary Figure S2) [29 30]. When not adjusting for age or sex, a total of 255 (14.53%) diseases are positively and significantly associated with PDI and 257 (14.64%) diseases are negatively and significantly associated with PDI. ‘Mental Disorders’ remains the category with the largest percent of diseases significantly and positively associated with PDI, with 55 out of 72 (76.39%) (Supplementary Figure S3 and Supplementary Table S6).

**Figure 3.**
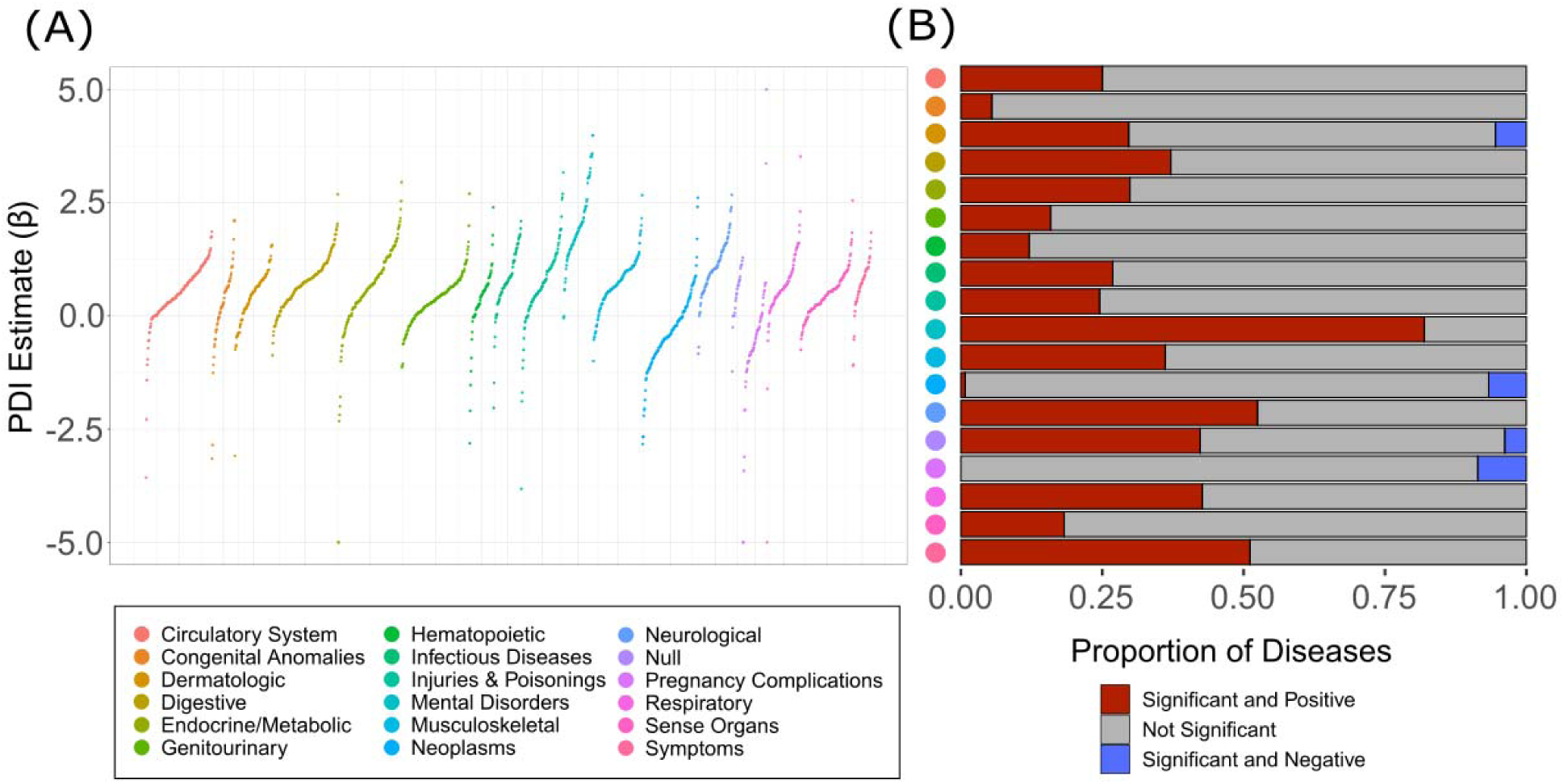
Phenome-wide associations between disease status and the perceived discrimination index (PDI). (A) Beta coefficients for PDI effect size estimates for associations with individual diseases are shown on the y-axis. Values greater than 5 or less than -5 were coerced to 5 and -5, respectively. Colors represent different disease categories, as indicated by the color key. (B) Proportion of diseases that show significant and positive (red), not significant (gray), and significant and negative associations with PDI.

**Table 1.** Diseases most strongly and positively associated with perceived discrimination index (PDI). Model specification: Disease ∼ PDI + age + sex.

| Phecode / Disease | Logistic regression model coefficients |  |  |  |
| --- | --- | --- | --- | --- |
|  | Estimate | SE | z-value | p-value |
| 300.9 / Posttraumatic stress disorder | 3.51 | 1.08e-1 | 32.50 | 1.19e-231 |
| 296.22 / Major depressive disorder | 1.94 | 6.50e-2 | 29.83 | 1.41e-195 |
| 296 / Mood disorders | 1.94 | 7.02e-2 | 27.70 | 7.66e-169 |
| 318 / Tobacco use disorder | 2.09 | 7.86e-2 | 26.63 | 3.19e-156 |
| 296.1 / Bipolar | 3.06 | 1.17e-1 | 26.08 | 5.83e-150 |
| 300.1 / Anxiety disorder | 1.48 | 6.29e-2 | 23.49 | 5.03e-122 |
| 316 / Substance addiction and disorders | 2.29 | 9.79e-2 | 23.42 | 2.65e-121 |
| 300 / Anxiety, phobic and dissociative disorders | 1.66 | 7.12e-2 | 23.29 | 4.90e-120 |
| 278.11 / Morbid obesity | 1.80 | 7.83e-2 | 23.05 | 1.46e-177 |
| 278.1 / Obesity | 1.32 | 5.93e-2 | 22.21 | 2.59e-109 |

### Health disparities

To identify health disparities as diseases for which disease burden differs across SIRE groups in the *All of Us* participant body, case and control status for 1,755 diseases were modeled as a function of SIRE with age and sex at birth as covariates (Figure 4). Using White as the reference SIRE group, these analyses revealed that membership to the Asian SIRE group was positively and significantly associated with 22 (1.25%) diseases and negatively and significantly associated with 150 (8.55%) diseases. Membership to the Black SIRE group was positively and significantly associated with 351 (20%) diseases and negatively and significantly associated with 89 (5.07%) diseases. Membership to the Hispanic SIRE group was positively and significantly associated with 132 (7.52%) diseases and negatively and significantly associated with 146 (8.32%) diseases. Glaucoma (β=9.62e-2, p=4.97e-23), intestinal disaccharide deficiencies (β=7.14e-1, p=9.55e-16), and viral hepatitis B (β=1.53, p=5.52e-12) are the diseases for which case status was the most highly and positively associated with membership to the Asian SIRE group. Hypertension (β=1.32, p=7.55e-255), essential hypertension (β=1.17, p=3.00e-247), and type 2 diabetes (β=1.17, p=2.86e-227) are the diseases for which case status was the most highly and positively associated with membership to the Black SIRE group. Type 2 diabetes (β=8.43e-1, p=5.53e-106), *H. pylori* infections (β=1.96, p=4.27e-92), and type 2 diabetes with ophthalmic manifestations (β=1.52, p=1.16e-81) are the diseases for which case status was the most highly and positively associated with membership to the Hispanic SIRE group (Table 2). Different subsets of diseases are identified when not controlling for either age or sex (Supplementary Figure 5 and Supplementary Tables S7 and S8).

**Figure 4.**
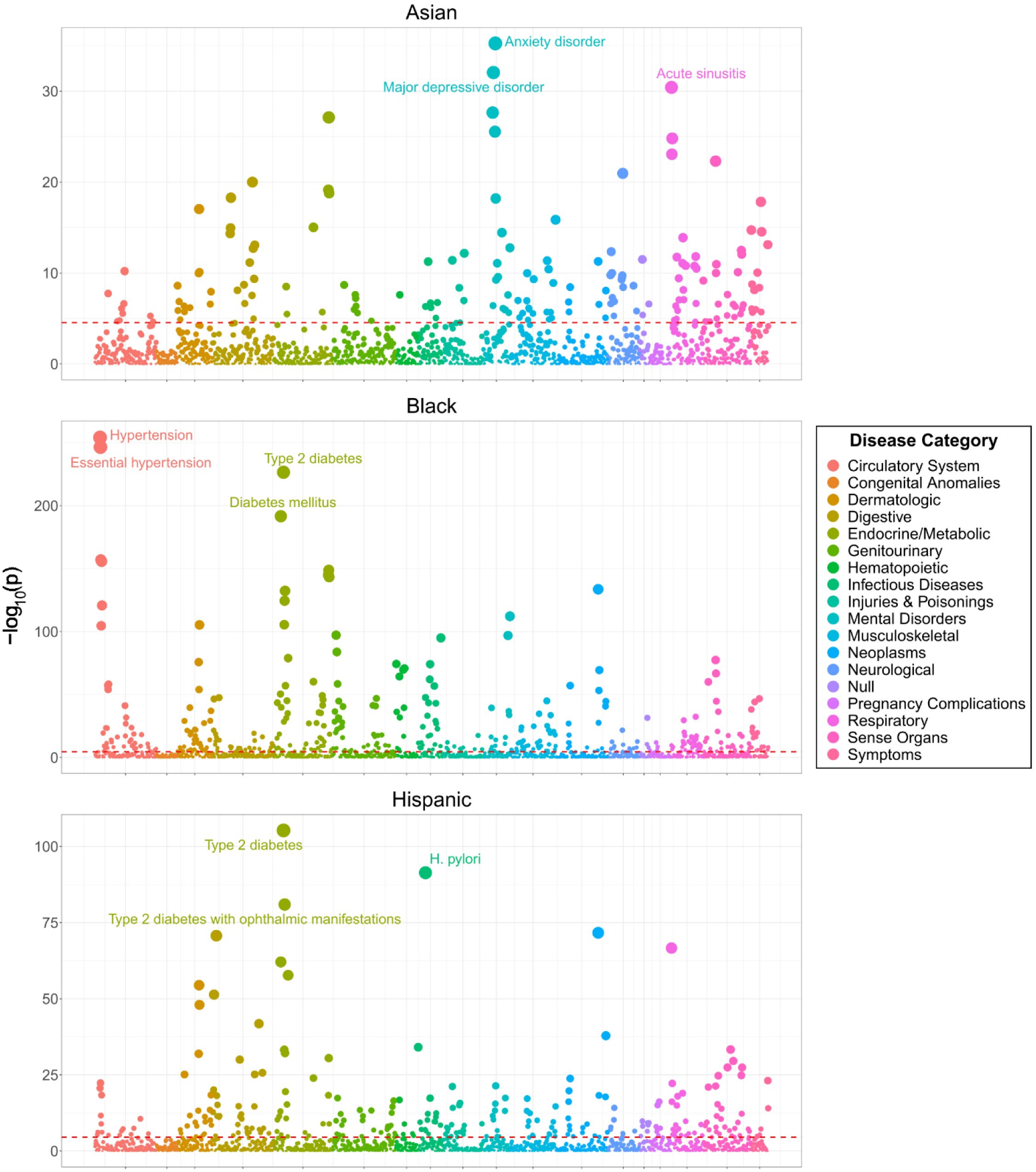
Phenome-wide associations between self-identified race and ethnicity (SIRE) and disease status. Model specification: Disease ∼ SIRE + age + sex. Points represent individual diseases. Point positions on y-axis represent -log_10_ p-value for the association between each disease-SIRE combination. Red lines represent the Bonferroni-adjusted -log_10_ p-value threshold of 4.545. Colors represent different disease categories, as indicated in the color key.

**Figure 5.**
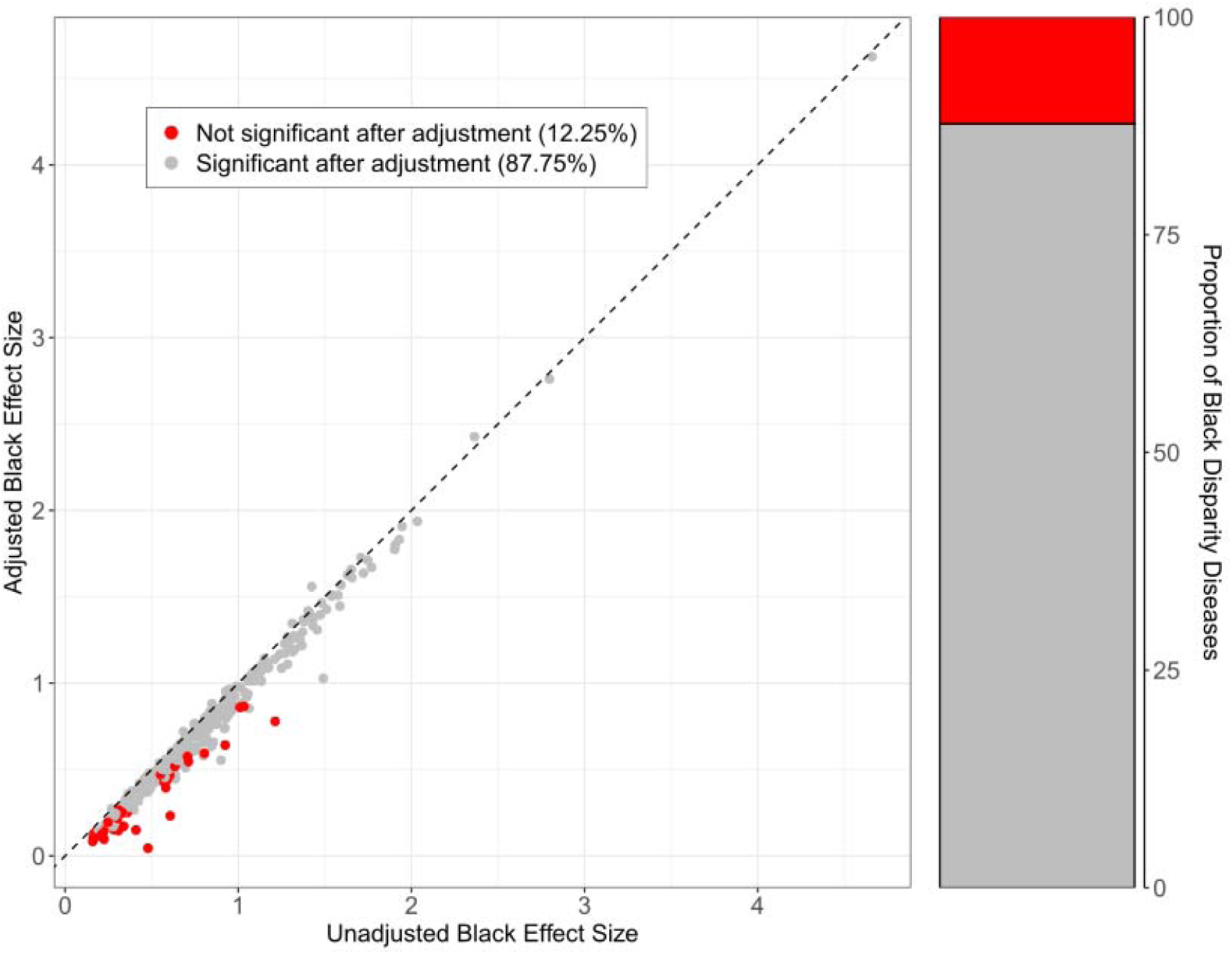
Association of Black SIRE with disease before (unadjusted) and after (adjusted) controlling for perceived discrimination (PDI). Unadjusted model specification: Disease ∼ SIRE + age + sex. Adjusted model specification: Disease ∼ SIRE + PDI + age + sex. Dashed line represents y = x. Points represent individual diseases and are colored by significance after adjustment for PDI: red (not significant after adjustment for PDI) and gra (remain significant after adjustment for PDI).

**Table 2.** Diseases most strongly and positively associated with Asian, Black, and Hispanic self-identified race and ethnicity (SIRE) groups. Model specification: Disease ∼ SIRE + age + sex.

|  | **Logistic regression model coefficients** |  |  |  |
| --- | --- | --- | --- | --- |
| **Phecode / Disease** | **Estimate** | **SE** | **z-value** | **p-value** |
| **Asian** |  |  |  |  |
| 365 / Glaucoma | 9.62e-1 | 9.74e-2 | 9.88e-0 | 4.97e-23 |
| 273.1 / Intestinal disaccharidase deficiencies and disaccharide malabsorption | 7.14e-1 | 8.89e-2 | 8.03e-0 | 9.55e-16 |
| 70.2 / Viral hepatitis B | 1.53e-0 | 2.22e-1 | 6.89e-0 | 5.52e-12 |
| 365.11 / Primary open angle glaucoma | 1.27e-0 | 1.87e-1 | 6.79e-0 | 1.08e-11 |
| 379.2/ Disorders of vitreous body | 6.32e-1 | 9.73e-2 | 6.50e-0 | 8.02e-11 |
| **Black** |  |  |  |  |
| 401 / Hypertension | 1.32e-0 | 3.88e-2 | 3.41e1 | 7.55e-255 |
| 401.1 / Essential hypertension | 1.17e-0 | 3.48e-2 | 3.36e1 | 3.00e-247 |
| 250.2 / Type 2 diabetes | 1.17e-0 | 3.63e-2 | 3.22e1 | 2.86e-227 |
| 250 / Diabetes mellitus | 1.24e-0 | 4.20e-2 | 2.96e1 | 2.45e-192 |
| 401.2 / Hypertensive heart and/or renal disease | 1.95e-0 | 7.27e-2 | 2.68e1 | 1.15e-157 |
| **Hispanic** |  |  |  |  |
| 250.2 / Type 2 Diabetes | 8.43e-1 | 3.86e-2 | 2.19e1 | 5.53e-106 |
| 41.8 / H. pylori | 1.96e-0 | 9.62e-2 | 2.04e1 | 4.27e-92 |
| 250.23 / Type 2 diabetes with ophthalmic manifestations | 1.52e-0 | 7.94e-2 | 1.91e1 | 1.16e-81 |
| 523 / Gingival and periodontal diseases | 1.15e-0 | 6.43e-2 | 1.79e1 | 2.04e-71 |
| 250 / Diabetes mellitus | 7.94e-1 | 4.75e-2 | 1.67e1 | 8.43e-63 |

### Perceived discrimination and health disparities

As described in the methods section, the contribution of perceived discrimination to SIRE disparities in disease burden was quantified with percent attenuation, PDI_attenuation_. Following this metric, perceived discrimination contributes the greatest to disparities in disease burden observed between the Black and White SIRE groups. The disease for which the greatest percent reduction was observed in the Black SIRE coefficient following adjustment for PDI is suicidal ideation (PDI_attenuation_=98.91%) followed by chronic pain syndrome (PDI_attenuation_=69.36%) and psychosis (PDI_attenuation_=64.38%). For each of these diseases, the Black SIRE coefficient was positive and significant in its association with disease status prior to PDI adjustment but was no longer significant following adjustment (Bonferroni adjusted p<1.42×10^-4^). This loss in significance was observed for a total of 43 (12.25%) out of the 351 diseases for which membership to the Black SIRE group was significantly and positively associated with disease status (Figure S5). The most common disease category represented by these diseases was ‘Digestive’, comprising 9 of the 43 (20.93%) diseases. This loss of significance was not observed in either the Asian or Hispanic SIRE groups, for whom values of PDI_attenuation_ were 6.08% or below for all diseases whose case status is positively and significantly associated with membership to either group (Table 3).

**Table 3.** Attenuation of associations between self-identified race and ethnicity (SIRE) and disease after adjustment by perceived discrimination index (PDI).

| Phocode / Disease | % reduction in estimate | Raw change in estimate | Significant after adjustment |
| --- | --- | --- | --- |
| **Asian** |  |  |  |
| 250.42 / Other abnormal glucose | 4.40% | -0.0160 | Yes |
| 366.2 / Senile cataract | 3.68% | -0.0172 | Yes |
| 365.11 / Primary open angle glaucoma | 3.68% | -0.0469 | Yes |
| 250.4 / Abnormal glucose | 3.66% | -0.0192 | Yes |
| 70.2 / Viral hepatitis B | 3.07% | -0.0458 | Yes |
| **Black** |  |  |  |
| 297.1 / Suicidal ideation | 98.91% | -0.47 | No |
| 355.1 / Chronic pain syndrome | 69.36% | -0.28 | No |
| 295.3 / Psychosis | 64.38% | -0.38 | No |
| 327.3 / Sleep apnea | 61.38% | -0.13 | No |
| 327.1 / Hypersomnia | 53.14% | -0.16 | No |
| **Hispanic** |  |  |  |
| 525 / Other diseases of the teeth and supporting structures | 6.08% | -0.0306 | Yes |
| 250.6 / Polyneuropathy in diabetes | 5.54% | -0.0330 | Yes |
| 70.3 / Viral hepatitis C | 5.32% | -0.0267 | Yes |
| 278.11 / Morbid obesity | 4.53% | -0.0123 | Yes |
| 401.21 / Hypertensive heart disease | 4.19% | -0.0236 | Yes |

These attenuation effects were further evaluated using mediation analyses, which were performed for each of the 43 diseases for which a loss of significance in the Black SIRE coefficient was observed after adjusting for PDI. The disease for which the indirect effect of PDI is the greatest is suicidal ideation, whose indirect effects of PDI constituted 97.54% of the total effects of PDI and SIRE on case status (Table 4). This is followed by chronic pain syndrome and sleep apnea, whose indirect effects accounted for 66.97% and 62.99% of their total effects, respectively. A strong correlation (Pearson’s r=0.99) is observed between the indirect effect of PDI from mediation analysis and PDI_attenuation_ when the former is expressed as a percentage of the total effect of membership to the Black SIRE group on case status (Supplementary Figure S6).

**Table 4.** Mediation analyses for Black-White disease disparities. The indirect effect of self-identified race and ethnicity (SIRE) on disease is mediated by perceived discrimination index (PDI). Results for are shown for diseases with the top 5 indirect effects.

| Phocode / Disease | Direct effect<sup>a</sup> | Indirect effect<sup>b</sup> | Total effect<sup>c</sup> | % indirect<sup>d</sup> |
| --- | --- | --- | --- | --- |
| 297.1 - Suicidal ideation | 1.12e-2<br>(1.12e-1) | 3.64e-1<br>(2.56e-2) | 3.73e-1<br>(1.02e-1) | 97.54 |
| 355.1 - Chronic pain syndrome | 1.20e-1<br>(9.01e-2) | 2.50e-1<br>(1.63e-2) | 3.74e-1<br>(8.55e-2) | 66.97 |
| 327.3 - Sleep apnea | 8.18e-2<br>(1.31e-1) | 1.37e-1<br>(2.60e-2) | 2.17e-1<br>(1.23e-1) | 62.99 |
| 295.3 - Psychosis | 2.09e-1<br>(4.90e-2) | 3.03e-1<br>(8.11e-3) | 5.10e-1<br>(4.78e-2) | 59.50 |
| 327.1 - Hypersomnia | 1.40e-1<br>(5.61e-2) | 1.57e-1<br>(1.33e-2) | 2.96e-1<br>(5.56e-2) | 53.22 |

<sup>a</sup> Direct of effect of SIRE on disease.
<sup>b</sup> Indirect of effect of SIRE on disease, mediated by PDI.
<sup>c</sup> Total effect of SIRE on disease: direct + indirect.
<sup>d</sup> Percent of total effect of SIRE on disease mediated by PDI.

## DISCUSSION

The PDI developed in this study provides a means of quantifying the degree of discrimination that *All of Us* participants experience. The composite metric captures most of the variance in participant responses to questions related to experiences of discrimination in the *All of Us* “Social Determinants of Health” survey. Furthermore, the metric is highly correlated with participant responses to each of the nine questions. These characteristics suggest that the PDI may be an effective means of allowing *All of Us* researchers to incorporate perceived discrimination as a variable in disease modeling.

The significant associations found between PDI values, SIRE, and disease status further underscore the potential of the PDI to be a proxy for perceived discrimination. Values for PDI tend to be higher for individuals identifying as either Asian or Black than those identifying as White. These results are consistent with those from prior studies on race and discrimination, which show that racial minority groups report more perceived discrimination than their White counterparts [22 23]. PDI was found to be positively and significantly associated with diseases from a wide range of disease categories. The disease category with the greatest proportion of positive associations with PDI is ‘Mental Disorders’. These results validate past research on discrimination and health, which have identified perceived discrimination as a potential predictor for a number of physical and especially mental disorders [5–12].

The results of the attenuation analyses performed provide evidence that discrimination contributes to racial disparities in disease burden in the *All of Us* participant body. This finding is reinforced by the accompanying mediation analyses performed, whose results also show that a number of SIRE-disease associations are partially attributable to perceived discrimination. However, while PDI values for both the Asian and Black SIRE groups were found to be significantly higher than the White SIRE group, perceived discrimination was found to only meaningfully contribute to health disparities between the Black and White SIRE groups. This may be a result of there being fewer health disparities between the Asian and White SIRE groups overall. Additionally, there may be other factors beyond perceived discrimination contributing to disparities between these groups.

While the results of the attenuation and mediation analyses performed may suggest that discrimination drives racial health disparities, the methods used are insufficient to discount the possibility of reverse causality. Trauma and depressive symptoms, such as those that occur in mental disorders such as depression and post-traumatic stress disorder, may lead to heightened perceptions of discrimination, as these diseases may impair an individual’s ability to properly assess threats [31 32]. More rigorous analyses are needed to elucidate the true nature of the PDI-disease associations observed in the *All of Us* participant body.

There are several important limitations to this study. As this is an observational study, there is the potential for unobserved confounding variables to influence study results. Other limitations are tied to the nature of the data collected by *All of Us*. As the *All of Us* participant body is comprised of volunteers, it is not a representative sample of the U.S. population. As a result, the participant body differs from the general population in several fundamental ways. The cohort has a greater prevalence of both common and rare diseases compared to the general population. Additionally, *All of Us* participants are more likely to be female, older, have more educational attainment, and identify as either Black or Hispanic than the general population [33]. Beyond limitations inherent to the study cohort, there are important caveats in the use of the Everyday Discrimination Scale on which the discrimination section in the *All of Us* “Social Determinants of Health” survey is based. It has been previously reported that the aspects of perceived discrimination that this scale captures may be limited, potentially necessitating the use of other discrimination measures in conjunction with this scale [34]. The findings reported here are therefore the most generalizable to the *All of Us* participant body and to the dimensions of discrimination captured by the Everyday Discrimination Scale.

These limitations notwithstanding, the methodology in calculating the PDI outlined here may support future discrimination-related research on the *All of Us* platform by providing a means through which perceived discrimination can be quantified. The associations between this metric and the case status for diseases spanning a variety of different categories validate the role of discrimination as an important SDOH. We also demonstrate the potential for feelings of perceived discrimination to contribute to racial disparities in disease burden on the *All of Us* platform. Efforts to mitigate racial health disparities may therefore benefit from addressing the disproportionate amount of discrimination suffered by socially disadvantaged groups.

## Data Availability

All code and data used in this study are made available on the All of Us Researcher Workbench.

## AUTHOR CONTRIBUTIONS

Vincent Lam and Sonali Gupta: data curation, methodology, analysis, visualization, and writing. I. King Jordan and Leonardo Mariño-Ramírez: Conceptualization, funding acquisition, methodology, project administration, visualization, and writing.

## SUPPLEMENTARY MATERIAL

Supplementary Material is available at JAMIA online

## ACKNOWLEDGEMENT

We gratefully acknowledge *All of Us* participants for their contributions, without whom this research would not have been possible. We also thank the National Institutes of Health’s All of Us Research Program for making available the participant data analyzed in this study.

## DATA AVAILABILITY

All code and data used in this study are made available on the All of Us Researcher Workbench.

