## Supplementary Materials for "Associations of perceived discrimination with health outcomes and health disparities in the *All of Us* cohort"

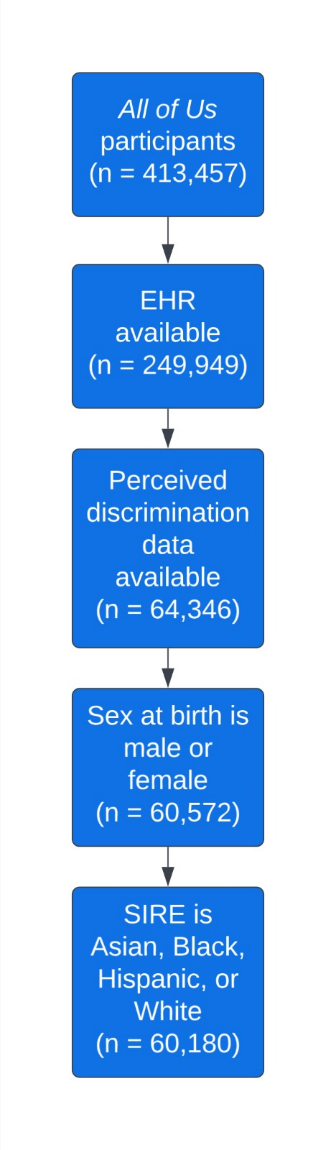

Figure S1. **All of Us participant cohort creation.** Inclusion criteria and the number of participants retained at each step.

### Table S1. **All of Us participant study cohort.**

| **Characteristic** | **Full Cohort** | **Asian** | **Black** | **Hispanic** | **White** |
| --- | --- | --- | --- | --- | --- |
| N participants | 60,180 (100%) | 1,291 (2.15%) | 4,726 (7.85%) | 5,336 (8.87%) | 48,827 (81.13%) |
| Mean age (SD) | 60.66 (15.81) | 62.23 (15.48) | 58.17 (13.77) | 50.87 (15.54) | 62.23 (15.48) |
| Female (%) | 39,118 (65.00) | 828 (64.14) | 3,498 (74.02%) | 3,844 (72.04%) | 30,948 (63.38) |
| Male (%) | 21,062 (35.00) | 463 (35.86) | 1,228 (25.98%) | 1,492 (27.96%) | 17,879 (36.62) |
| Mean PDI (SD) | 0.15 (0.16) | 0.19 (0.15) | 0.24 (0.21) | 0.18 (0.18) | 0.14 (0.15) |

### Table S2. **Discrimination questions in the “Social Determinants of Health” survey.**

| **Question** | **Text (“In your day-to-day life, how often [are you] …”)** |
| --- | --- |
| 1 | “…called names or insulted?” |
| 2 | “…threatened or harassed?” |
| 3 | “…treated with less courtesy than other people?” |
| 4 | “…treated with less respect than other people?” |
| 5 | “…people act as if they are afraid of you?” |
| 6 | “…people act as if they think you are dishonest?” |
| 7 | “…people act as if they think you are not smart?” |
| 8 | “…people act as if they’re better than you are?” |
| 9 | “…receive poorer service than other people at restaurants or stores?” |

Table S3. **Associations of self-identified race ethnicity (SIRE) with perceived discrimination index (PDI), controlling for age and sex.** Model specification: PDI ~ SIRE + age + sex.

|  | **Logistic regression model coefficients** | | | |
| --- | --- | --- | --- | --- |
| **SIRE** | **Estimate** | **SE** | **z-value** | **p-value** |
| Asian | 1.84e-2 | 4.27e-3 | 4.31 | 1.61e-5 |
| Black | 8.74e-2 | 2.30e-3 | 38.01 | 1.91e-312 |
| Hispanic | 3.59e-2 | 2.22e-3 | 1.62 | 1.05e-1 |

### Table S4. **Associations of self-identified race ethnicity (SIRE) with perceived discrimination index (PDI), not adjusted for age and sex: PDI ~ SIRE.**

|  | **Logistic regression model coefficients** | | | |
| --- | --- | --- | --- | --- |
| **SIRE** | **Estimate** | **SE** | **t-value** | **p-value** |
| Asian | 5.03e-2 | 4.41e-3 | 11.41 | 3.87e-30 |
| Black | 9.88e-2 | 2.38e-3 | 41.47 | 0.00 |
| Hispanic | 3.54e-2 | 2.26e-3 | 15.70 | 1.91e-55 |

### Table S5. **Associations between reasons for participant experiences of discrimination and the perceived discrimination index (PDI): PDI ~ Reasons.**

|  | **Logistic regression model coefficients** | | | |
| --- | --- | --- | --- | --- |
| **Attribution** (Question text: “What do you think is the main reason for these experiences?” | **Estimate** | **SE** | **t-value** | **p-value** |
| Some Other Aspect of Your Physical Appearance | 7.33e-2 | 2.44e-3 | 30.00 | 5.76e-195 |
| Your Weight | 5.75e-2 | 2.19e-3 | 26.27 | 1.87e-150 |
| Your Race | 5.08e-2 | 2.09e-3 | 24.23 | 1.49e-128 |
| Your Sexual Orientation | 6.57e-2 | 3.71e-3 | 17.71 | 7.13e-70 |
| Your Ancestry or National Origins | 4.34e-2 | 2.56e-3 | 16.94 | 4.53e-64 |
| Your Gender | 3.19e-2 | 2.34e-3 | 13.62 | 3.85e-42 |
| Your Education | 2.65e-2 | 2.28e-3 | 11.64 | 2.97e-31 |
| Your Religion | 3.93e-2 | 3.62e-3 | 10.84 | 2.49e-27 |
| Your Height | 3.09e-2 | 3.15e-3 | 9.83 | 9.46e-23 |
| Your Age | -1.68e-2 | 1.74e-3 | -9.60 | 8.93e-22 |
| Other (specify) | 1.56e-2 | 1.75e-3 | 8.91 | 5.39e-19 |

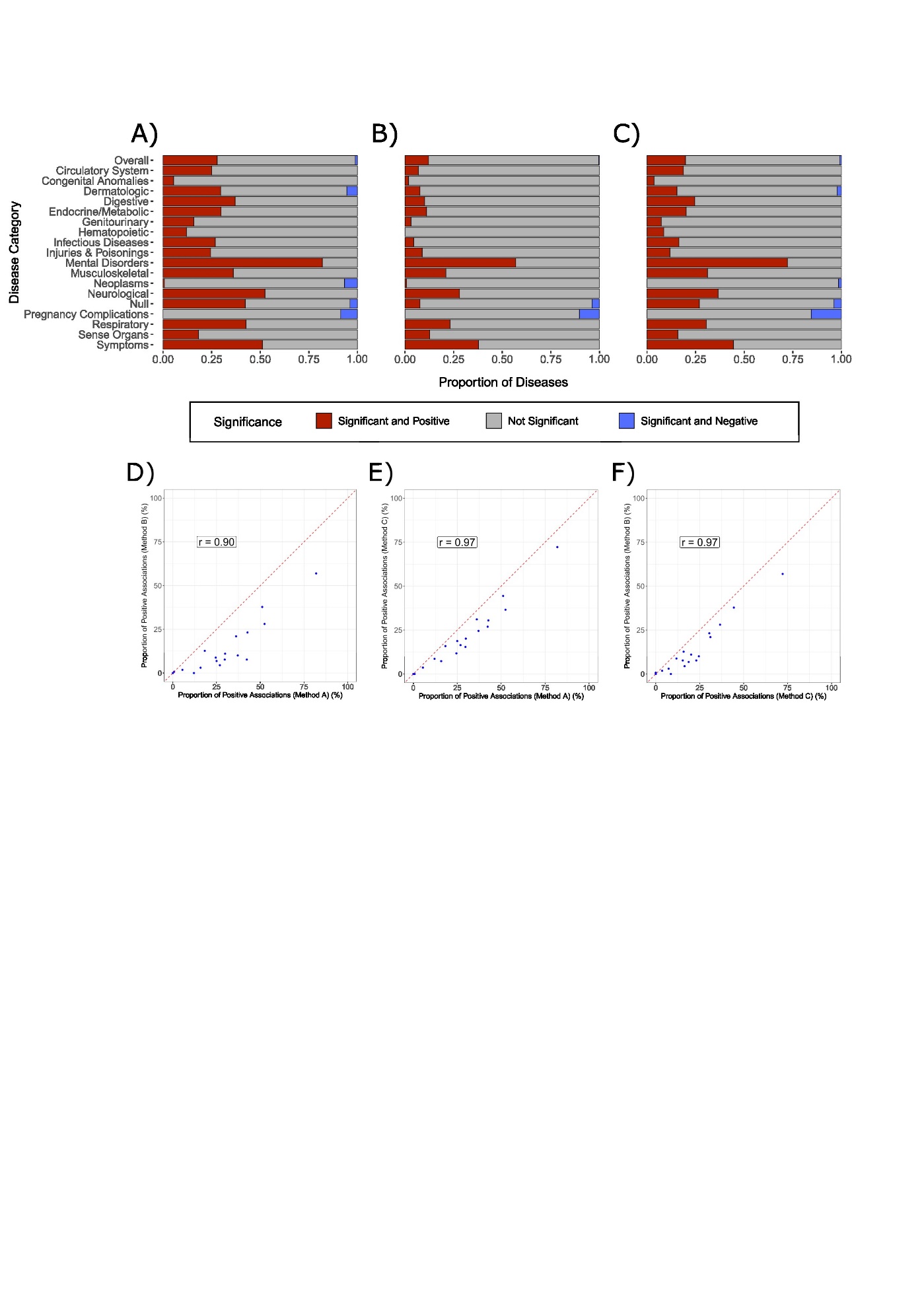

Figure S2. **Evaluation of different methods of measuring perceived discrimination.** Method A is the use of the continuous PDI metric. Methods B and C entail the use ­­ labels, whereby participants are classified as having ‘low’, ‘moderate’, and ‘high’ levels of perceived discrimination based on pre-established thresholds. In Method B, participant responses to each of the nine discrimination questions in the “Social Determinants of Health” survey were converted to ordinal values ranging from 0-5 using the scheme described in the manuscript. For each participant, the mean ordinal value across all 9 domains was calculated. Participants were then assigned a label of ‘low’, ‘medium’, or ‘high’ based on terciles of the means of All of Us participants who responded to the discrimination questions, following the scheme employed by Forde et al. In Method C, participants were assigned labels of ‘low’, ‘medium’, and ‘high’ based on thresholds informed by distributions of PDI for all participants for whom PDI could be computed. The thresholds for ‘low’, ‘medium’, and ‘high’ are $0-0.\bar{6}$, $0.\bar{6}-0.2\bar{3}$, and $0.2\bar{3}-1$, respectively. Panels A, B, and C show disease categories and their associations with PDI, using Methods A, B, and C to measure discrimination, respectively (Disease ~ Discrimination + age + sex). Red bars represent the proportion of diseases in each category significantly and positively associated with PDI, blue the proportion negatively and significantly associated with PDI, and gray the proportion not significantly associated with PDI, as indicated by the figure legend. Panels D, E, and F show pairwise comparisons between each of the three methods used to measure discrimination in the form of regression plots. Pearson correlation scores are shown. Points represent individual disease categories, and the red dashed line represents x = y. Values on either axis represent the proportion of diseases in the category that are positively and significantly associated with discrimination using the given method of measuring discrimination.

| 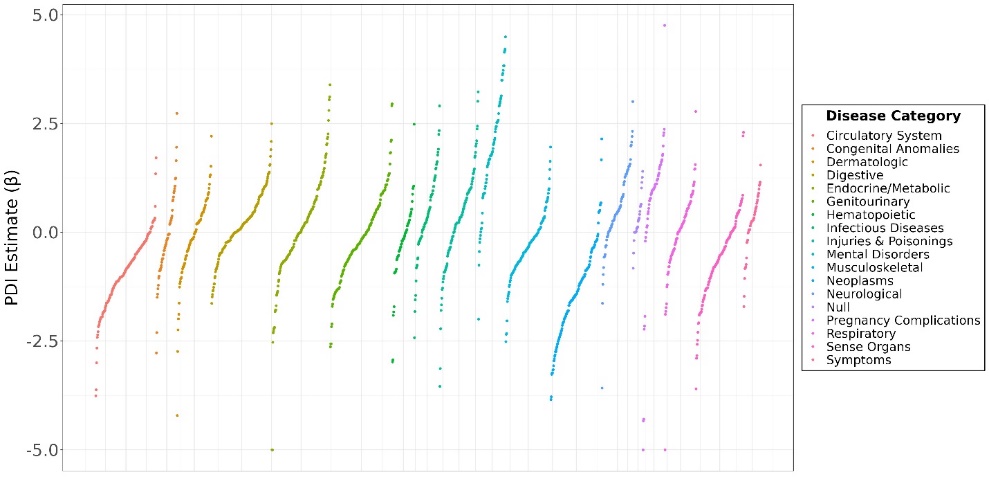 | 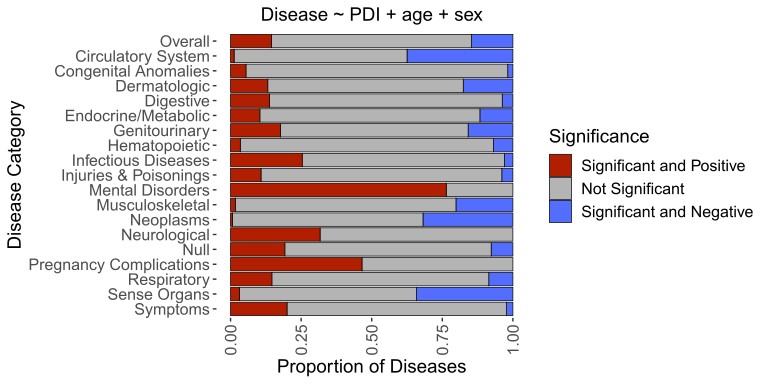 |
| --- | --- |

Figure S3. **Phenome-wide associations between disease status and the perceived discrimination index (PDI).** Not adjusted for age and sex. (A) Beta coefficients for PDI effect size estimates for associations with individual diseases are shown on the y-axis. Values greater than 5 or less than -5 were coerced to 5 and -5, respectively. Colors represent different disease categories, as indicated by the color key. (B) Proportion of diseases that show significant and positive (red), not significant (gray), and significant and negative associations with PDI.

Table S6. **Diseases most strongly and positively associated with perceived discrimination index PDI.** Not adjusted for age or sex. Status ~ PDI.

|  | **Logistic regression model coefficients** | | | |
| --- | --- | --- | --- | --- |
| **Phecode / Disease** | **Estimate** | **SE** | **t-value** | **p -value** |
| 300.9 / Posttraumatic stress disorder | 3.72 | 1.03e-1 | 36.13 | 8.69e-286 |
| 296.22 / Major depressive disorder | 2.07 | 6.20e-2 | 33.37 | 3.79e-244 |
| 296.1 / Bipolar | 3.50 | 1.11e-1 | 31.52 | 5.32e-216 |
| 300.1 / Anxiety disorder | 1.77 | 6.00e-2 | 29.46 | 8.16e-191 |
| 296 / Mood disorders | 1.86 | 6.65e-2 | 27.92 | 1.37e-171 |
| 297.1 / Suicidal ideation | 4.14 | 1.52e-1 | 27.26 | 1.19e-163 |
| 318 / Tobacco use disorder | 2.02 | 7.48e-2 | 26.95 | 5.76e-160 |
| 316 / Substance addiction and disorders | 2.42 | 9.33e-2 | 25.94 | 2.10e-148 |
| 300.11 / Generalized anxiety disorders | 2.16 | 8.35e-2 | 25.92 | 4.43e-148 |
| 300 / Anxiety, phobic, and dissociative disorders | 1.66 | 6.77e-2 | 24.56 | 3.71e-133 |

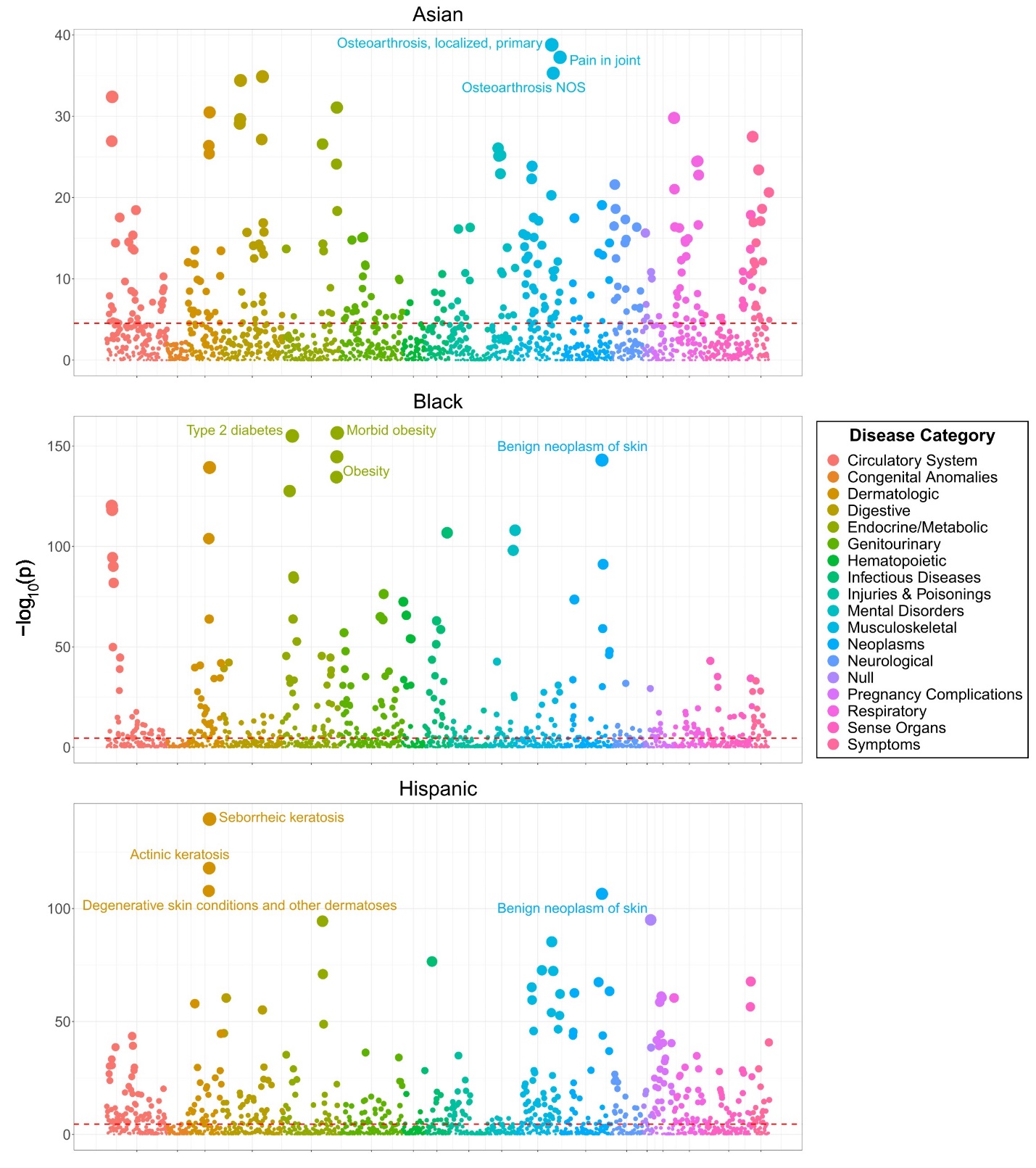

Figure S5. **Phenome-wide associations between self-identified race and ethnicity (SIRE) and disease status.** Not adjusted for age and sex. Model specification: Disease ~ SIRE. Points represent individual diseases. Point positions on y-axis represent -log_10_ p-value for the association between each disease-SIRE combination. Red lines represent the Bonferroni-adjusted -log_10_ p-value threshold of 4.545. Colors represent different disease categories, as indicated in the color key.

Table S7. **Diseases most strongly and positively associated with membership to the Asian, Black, and Hispanic self-identified race and ethnicity (SIRE) groups.** Not adjusted for age or sex. Model specification: Disease ~ SIRE.

|  | **Logistic regression model coefficients** | | | |
| --- | --- | --- | --- | --- |
| **Phecode / Disease** | **Estimate** | **SE** | **t-value** | **p-value** |
| **Asian** | | | | |
| 70.2 / Viral hepatitis B | 1.27 | 2.17e-1 | 5.87 | 4.47e-9 |
| 282.8 / Other hemoglobinopathies | 1.08 | 2.24e-1 | 4.83 | 1.37e-6 |
| 271.3 / Intestinal disaccharidase deficiencies and disaccharide malabsorption | 4.18e-1 | 8.72e-2 | 4.79 | 1.63e-6 |
| 360.2 / Progressive myopia | 1.07 | 2.29e-1 | 4.68 | 2.88e-6 |
| 644 / Anemia during pregnancy | 0.96 | 2.09e-1 | 4.62 | 3.86e-6 |
| **Black** | | | | |
| 278.11 / Morbid obesity | 1.06 | 3.96e-2 | 26.71 | 3.29e-157 |
| 250.2 / Type 2 diabetes | 9.22e-1 | 3.47e-2 | 26.59 | 9.90e-156 |
| 278.1 / Obesity | 8.11e-1 | 3.16e-2 | 25.67 | 2.46e-145 |
| 278 / Overweight, obesity and other hyperalimentation | 8.89e-1 | 3.59e-2 | 24.75 | 3.22e-135 |
| 250 / Diabetes mellitus | 9.60e-1 | 3.98e-2 | 24.10 | 2.71e-128 |
| **Hispanic** | | | | |
| 1010.6 / (Unnamed Disease) | 1.14 | 5.49e-2 | 20.76 | 1.03e-95 |
| 41.8 / *H. pylori* | 1.69 | 9.10e-2 | 18.61 | 2.77e-77 |
| 649 / Other conditions or status of the mother complication pregnancy, childbirth, or the puerperium | 9.96e-1 | 6.00e-2 | 16.61 | 6.31e-62 |
| 650 / Normal delivery | 1.15 | 6.99e-2 | 16.53 | 2.33e-61 |
| 523 / Gingival and periodontal diseases | 1.01 | 6.12e-2 | 16.50 | 3.86e-61 |

Table S8. **Counts of diseases significantly associated with minority self-identified race and ethnicity (SIRE) groups.** Not adjusted for age or sex. Model specification: Disease ~ SIRE.

| **SIRE** | **# of diseases with a positive and significant association (%)** | **# of diseases with a negative and significant association (%)** | **Disease categories with greatest proportion of diseases represented among positively associated diseases** |
| --- | --- | --- | --- |
| Asian | 7 | 282 | Null; 1/26 (3.85%) |
| Black | 297 | 126 | Infectious Diseases; 24 / 67 (35.8%) |
| Hispanic | 103 | 416 | Pregnancy Complications; 32 / 58 (55.17%) |

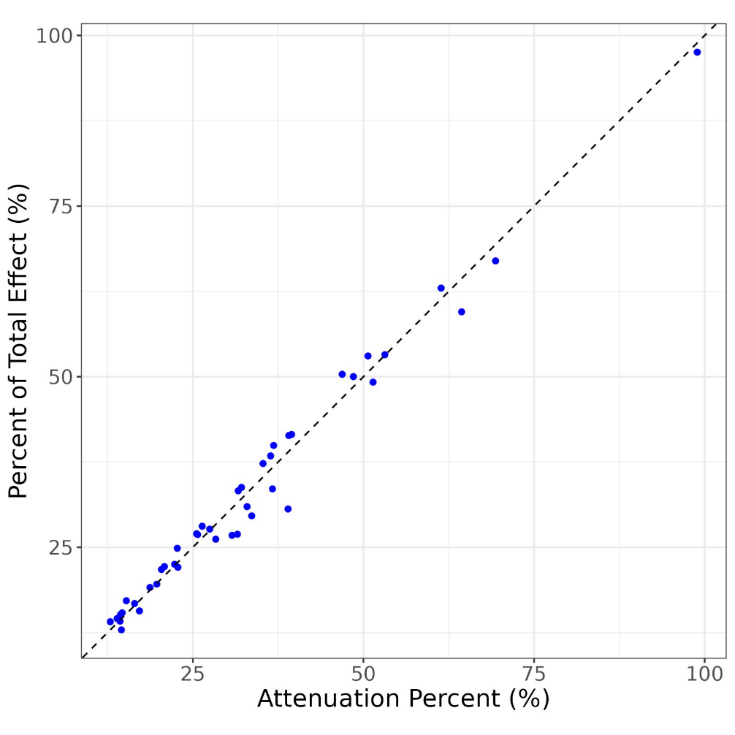

Figure S6. **Comparison of mediation and attenuation analysis for the effect of perceived discrimination index (PDI) on Black-White health disparities.** Percent of total effect on disease status comprised of the indirect effect of PDI from mediation analysis (y-axis). Percent attenuation of Black SIRE coefficient following adjustment for PDI. Points represent individual diseases. Dashed line represents y = x.
